# Clinical predictors for etiology of acute diarrhea in children in resource-limited settings

**DOI:** 10.1101/2020.01.27.20016725

**Authors:** Benjamin Brintz, Joel Howard, Benjamin Haaland, James A. Platts-Mills, Tom Greene, Adam C. Levine, Eric Nelson, Andrew T. Pavia, Karen L. Kotloff, Daniel T. Leung

## Abstract

**Background:** Diarrhea is one of the leading causes of childhood morbidity and mortality in lower- and middle-income countries. In such settings, access to laboratory diagnostics are often limited, and decisions for use of antimicrobials often empiric. Clinical predictors are a potential non-laboratory method to more accurately assess diarrheal etiology, the knowledge of which could improve management of pediatric diarrhea.

**Methods:** We used clinical and quantitative molecular etiologic data from the Global Enteric Multicenter Study (GEMS), a prospective, case-control study, to develop predictive models for the etiology of diarrhea. Using random forests, we screened the available variables and then assessed the performance of predictions from random forest regression models and logistic regression models using 5-fold cross-validation.

**Results:** We identified 1049 cases where a virus was the only etiology, and developed predictive models against 2317 cases where the etiology was known but non-viral (bacterial, protozoal, or mixed). Variables predictive of a viral etiology included lower age, a dry and cold season, increased height-for-age z-score (HAZ), lack of bloody diarrhea, and presence of vomiting. Cross-validation suggests an AUC of 0.825 can be achieved with a parsimonious model of 5 variables, achieving a specificity of 0.85, a sensitivity of 0.59, a NPV of 0.82 and a PPV of 0.64.

**Conclusion:** Predictors of the etiology of pediatric diarrhea can be used by providers in low-resources setting to inform clinical decision-making. The use of non-laboratory methods to diagnose viral causes of diarrhea could be a step towards reducing inappropriate antibiotic prescription worldwide.

**Author Summary:** Diarrhea is one of the leading causes of death in young children worldwide. In low-resource settings, diarrhea testing is not available or too expensive, and the decision to prescribe antibiotics is often made without testing. Using clinical information to predict which cases are caused by viruses, and thus wouldn’t need antibiotics, would help to improve appropriate use of antibiotics. We used data from a large study of childhood diarrhea, paired with advanced statistical methods including machine learning, to come up with the top clinical factors that could predict a viral cause of diarrhea. We compared 1049 cases where a virus was the only cause, with 2317 cases where the cause was known but not a virus. We found that a lower age, dry and cold season, nutritional status defined by increased height, lack of blood diarrhea, and vomiting, were the clinical factors most predictive of whether the diarrhea was caused by a virus. We found that, using just those 5 factors, we were able to predict a viral cause with good accuracy. Our findings can be used by doctors to guide the appropriate use of antibiotics for diarrhea in children.

## Introduction

Diarrhea is one of the leading causes of childhood morbidity and mortality in lower- and middle-income countries (LMICs) and is among the most common reasons for admission into a health facility [1]. Treatment of diarrhea is commonly empiric, with antibiotic prescription mostly based on clinical suspicion of bacterial etiology, such as in cases of bloody diarrhea. In resource-limited settings, laboratory etiological diagnosis is rarely made due to cost constraints or availability. Despite Integrated Management of Childhood Illness (IMCI) guidelines recommending use of antibiotics only for cases of bloody diarrhea and suspected cholera, studies have demonstrated that over 42% of young children with non-bloody diarrhea receive antibiotics, with the rate of use varying widely by country and setting [2]. This inappropriate use of antimicrobials can lead to toxicity, increased costs of care, and development of resistance [3]. Additionally, previous studies predicting etiology of diarrheal illness have been limited by the low number of participants, a lack of controls without diarrhea, single center design, and insufficient stool testing [14-17]. Thus, methods providing clinical decision support that accurately predict diarrhea etiology and reduce reliance on laboratory testing are needed. Recently, tools for decision making and clinical prediction have been bolstered by the accessibility of machine learning methods such as random forests, neural networks, and support vector machines [4].

The availability of molecular diagnostics in recent years has enabled accurate determination of etiology for pediatric diarrhea. In several large studies in LMICs, this has been used for estimating the population-based burden of various diarrheal pathogens [5-7]. While etiologies of diarrhea are now better-understood, there remains a gap in knowledge regarding clinical predictors for improving clinical decision making in the setting of infectious diarrhea. In this study, we use data from the Global Enteric Multicenter Study (GEMS) [5] to examine clinical diagnostic predictors of diarrhea etiology.

## Methods

### Study Design and Settings

GEMS is a prospective, case-control study that took place from 2007-2011 in 7 countries in Africa and South Asia (Figure S2). There were 9439 children with moderate-to-severe diarrhea (MSD) enrolled at local health care centers along with 1 to 3 matched non-diarrheal controls. An acute episode of diarrhea was defined as MSD if it had onset within the past 7 days and fulfilled at least one of the following criteria: sunken eyes, more than normal; loss of skin turgor; intravenous hydration administered or prescribed; visible blood in stool or parental report; or admission to hospital with diarrhea or dysentery. At enrollment, a stool sample was taken from each child to identify enteropathogens along with clinical information, including demographic, anthropometric, and clinical history. Methods for GEMS have been described in detail previously [5, 8, 9]. Because pathogen nucleic acids are frequently detected by PCR in children without diarrhea, we used the quantitative real-time PCR-based (qPCR) majority attribution models developed by Liu et al [6] to assign etiology of diarrhea. We derived site- and age-specific attributable fractions (AFe) for each episode, and used a cut-off of greater than 0.5 to indicate attribution of a pathogen to a particular episode. We defined viral etiology as majority attribution of the diarrhea episode by viral pathogen(s) only (i.e. excluding any co-infections with bacteria or protozoa). We defined other known etiologies as having a majority attribution of diarrhea episode by at least one other non-viral pathogen. Additionally, we defined a bacterial etiology as attribution of the diarrhea episode by any bacterial pathogen, including cases in which more than one pathogen was attributed (i.e. bacteria and virus, or bacteria and protozoa, or multiple bacteria). For patients with unknown etiologies, we presume there is an infectious cause to their diarrhea that we are not detecting, and excluded these cases from our predictive model.

We used the patient’s clinical symptoms data, epidemiologic, and anthropometric data at presentation as potential predictors of etiology. We used standard guidelines from the transparent reporting of a multivariable prediction model for individual diagnosis (TRIPOD) to develop our prediction model [10]. We focused on the prediction of a viral etiology of acute diarrhea versus all other known etiologies as knowing this could offer support for providers to withhold antibiotics. We additionally looked at the prediction of any bacterial pathogen as a way to determine if follow-up testing, such as stool culture for antimicrobial agent susceptibilities, may be helpful in ambiguous cases.

### Data Processing

We performed all data processing and analyses using R version 3.6.2 [11]. Starting with over 1000 variables collected, we excluded all variables which would not be available at the time of presentation. Questions which had very few responses in certain categories (<10) were regrouped into an “other” category as appropriate. 3 patients responded they “Don’t Know” when asked if they had any blood in their stool since the illness began and were removed from the dataset. There were 43 patients with other forms of missing data which were additionally removed for a total of 46 patients removed out of 3412. We maximized the utility of the modeling process by removing highly collinear and similar variables (e.g. weight-, BMI, and BMI-for-age z-scores). These steps left 156 potential predictor variables for analysis.

In addition to the information from the GEMS survey, we developed a season variable using temperature and rain information from NOAA weather stations close to the health centers and with data during the GEMS time period [12]. We defined a rainy season day as a day having a center-aligned 1-month moving rain average greater than the overall rain average within the study period. We defined a hot season day as a day having a center-aligned 1-month moving temperature average greater than the overall temperature average within the study period. The season variable was an indicator for a rainy/hot day, rainy/cold day, dry/cold day, or dry/hot day.

### Statistical Modeling and Assessment

We used random forests as a screening step to obtain an order of variable importance toward the goal of building a parsimonious model. The random forest method uses an ensemble approach by generating multiple decision trees (1000 trees, square root of the number of predictors considered by each tree when splitting a node (12)) and assesses variable importance by determining a reduction in mean squared prediction error for each variable on the “out-of-bag” samples (or testing samples) created while bootstrapping the data. We used random forests for variable selection in order to determine if there might be some complexity (non-linearity or interactions) in the predictors that could not be explained by an additive model. During this step, categorical variables are treated as a single variable with an indicator for each categorical level. We additionally test for robustness of this variable importance measure by varying the numbers of trees and predictors considered per node split.

We used 5-fold cross-validation to attain an estimate of generalizable model performance. For each cross-validation iteration (100 total), we re-fit the random forest regression described above to get an order of variable importance for each training set to determine which variables we used to fit separate logistic regression, random forest, gradient boosted regression trees and vanilla neural network models with various predictor subset sizes. Subsets examined were sizes 1 through 10, 15, 20, 30, 40, and 50. Tree based models used 1000 trees, and we chose to use twice as many nodes as the number of predictors in the neural network’s hidden layer. In each iteration of cross-validation we made predictions on the test set and obtained measures of performance: the receiver operating characteristic (ROC) curve, and area under the ROC curve (AUC), also known as the C-statistic, along with AUC 95% confidence intervals [13]. For a diagnostic threshold balancing the relative costs of false positives and false negatives, we calculated the positive predictive value (PPV) and the negative predictive value (NPV) as functions of the derived sensitivity and specificity of the prediction, using the prevalence of the corresponding etiology in GEMS. Finally, from the cross-validation, we determined how calibrated the different size models were by comparing each predicted probability of viral age (x-axis) with the observed proportion of viral cases within 0.05 plus or minus the predicted probability (y-axis) and report the intercept (Steyerberg’s A) and slope (Steyerberg’s B) of a fitted simple linear regression model [27]. In order to assess the robustness of the model and variable importance, we observe site-specific variable importance, look at site- and continent-specific cross-validated AUCs, and perform a leave-one-site-out pseudo external-validation.

### Ethics approval

The GEMS study protocol was approved by ethics committees at the University of Maryland, Baltimore and at each field site. Parents or caregivers of participants provided written informed consent, and a witnessed consent was obtained for illiterate parents or caretakers.

## Results

Of the 9439 patients in the GEMS study with MSD, 3366 are included in this analysis (Figure S3), 1049 had a viral etiology and 2069 had a bacterial etiology (Table 1). Using random forest screening, we found that age, season, bloody diarrhea, height-for-age z-score (HAZ), and vomiting were the five variables most predictive of a viral etiology (Table 2), and that top predictive variables for bacterial etiology were similar (Supplemental Table S2). The top five predictors did not change order with the number of trees increased or the number of predictors per split set at 6, 16, or 25. All predictors considered are shown in Supplemental Table 3 (survey variable names available at https://github.com/LeungLab/GEMSClinicalPredictors/).

**Table 1:** Number of cases attributed to each pathogen with an attributable fraction above 0.5.

| Pathogen | Cases |
| --- | --- |
| *Adenovirus 40/41* | 222 |
| *Aeromonas* | 59 |
| *Astrovirus* | 111 |
| *C. jejuni/C. coli* | 85 |
| *Cryptosporidium* | 301 |
| *Cyclospora cayetanensis* | 16 |
| *Entamoeba histolytica* | 29 |
| *Helicobacter pylori* | 131 |
| *Isospora* | 3 |
| *Norovirus GII* | 70 |
| *Rotavirus* | 967 |
| *Salmonella* | 67 |
| *Sapovirus* | 75 |
| *Shigella/EIEC* | 1376 |
| *Vibrio cholerae* | 152 |
| *EAEC* | 1 |
| *ST-ETEC (STh)* | 407 |
| *Typical EPEC (bfpA)* | 43 |
| Occurrences | Cases |
| *Protozoal* | 218 |
| *Viral* | 1049 |
| *Viral-Protozoal* | 30 |
| *Bacterial* | 1664 |
| *Bacterial-Protozoal* | 92 |
| *Bacterial-Viral* | 307 |
| *Bacterial-Viral-Protozoal* | 6 |

**Table 2:** Rank of variable importance by reduction in residual sum of squares (RSS) using random forest regression.

| **Viral Etiology** |  |
| --- | --- |
| **Variable Name** | **RSS Reduction** |
| Age | 51.6 |
| Season | 29.0 |
| Blood in stool | 26.1 |
| HAZ | 24.7 |
| Vomiting | 23.0 |
| Breastfed | 22.0 |
| MUAC | 20.9 |
| Resp. Rate | 18.5 |
| Wealth Index | 18.3 |
| Temperature | 16.7 |

When we performed 5-fold cross-validated logistic regression and random forest models, the average AUC across 100 random iterations of cross-validation ranged from 0.71 (1 variable) to 0.84 (8 or more variables) for prediction of viral etiology (Figure 1) with similar results for bacterial etiology (Figure S4). Although the neural network outperforms the logistic regression by about 0.5% AUC at a smaller number of variables, we determined the gradient boost regression trees and neural network models did not improve discrimination beyond their simpler counterparts enough to pursue them further in this context (Figure S5). Our method for assessing calibration showed that the logistic regression model was better calibrated than the random forest model with more than 1 variable included and that models between 3 and 15 variables were similarly well-calibrated (Table S4). We demonstrate the direction and magnitude of the effect of the top 10 variables from variable importance screening by fitting a logistic regression on the entire data set (Table 3) and by generating partial dependency plots from the random forest regression (Figure S6). We additionally include the logistic regression coefficients for the top 5 variable model in the supplement (Table S5) as well as compare the distribution of predictions for our 3366 cases versus the 1892 cases with qPCR data but no etiology defined (Figure S7). Lower age, a higher HAZ, more vomiting, no blood in the stool, and a dry/cold season, were associated with viral etiology. As expected, the opposite associations were found for bacterial etiology (Supplemental Table S6). We found similar results in a sensitivity analysis with rotavirus removed (for generalization of these results to locations with high rotavirus vaccine coverage), though some effect magnitudes were reduced (Table S7). Given the similarity of the results between the logistic regression models and random forest regression models, we conducted all successive analyses using the simpler logistic regression. To estimate the achievable sensitivity and specificity by each model at various predictor sizes, we generated ROC curves from cross-validation, and found that using a parsimonious model of 5 variables, we achieved a specificity of 0.85 and a sensitivity of close to 0.60 for prediction of viral etiology (Figure 2). For predicting a bacterial cause, our models achieved a sensitivity of 0.85 and a specificity of 0.63 (Figure S8). Using the prevalence of viral etiology in GEMS, our prediction model had a NPV of 0.82 and a PPV of 0.64.

**Table 3:** The odds ratios, 95% confidence interval, and p-value from a logistic regression model for the viral only outcome.

| Variable Name | Odds Ratios (95% CI) | P-value |
| --- | --- | --- |
| Intercept | 1.975 (0.053 – 72.894) | 0.7117 |
| Age (mo.) | 0.956 (0.944 – 0.967) | <0.0001 |
| Season |  |  |
| Dry/Cold | Reference |  |
| Rainy/Cold | 0.197 (0.145 – 0.268) | <0.0001 |
| Dry/Hot | 0.304 (0.244 – 0.379) | <0.0001 |
| Rainy/Hot | 0.338 (0.268 – 0.426) | <0.0001 |
| Blood in stool | 0.129 (0.096 – 0.173) | <0.0001 |
| HAZ | 1.168 (1.081 – 1.262) | 0.0001 |
| Vomiting | 2.383 (1.995 – 2.847) | <0.0001 |
| Breastfed |  |  |
| None | Reference |  |
| Partially | 2.359 (1.827 – 3.046) | <0.0001 |
| Exclusively | 2.400 (1.554 – 3.705) | 0.0001 |
| MUAC | 1.031 (0.963 – 1.105) | 0.3773 |
| Resp. Rate (per min.) | 0.990 (0.979 – 1.000) | 0.0541 |
| Wealth Index | 1.066 (0.976 – 1.164) | 0.1559 |
| Temperature (°C) | 0.988 (0.897 – 1.088) | 0.8022 |

**Figure 1:**
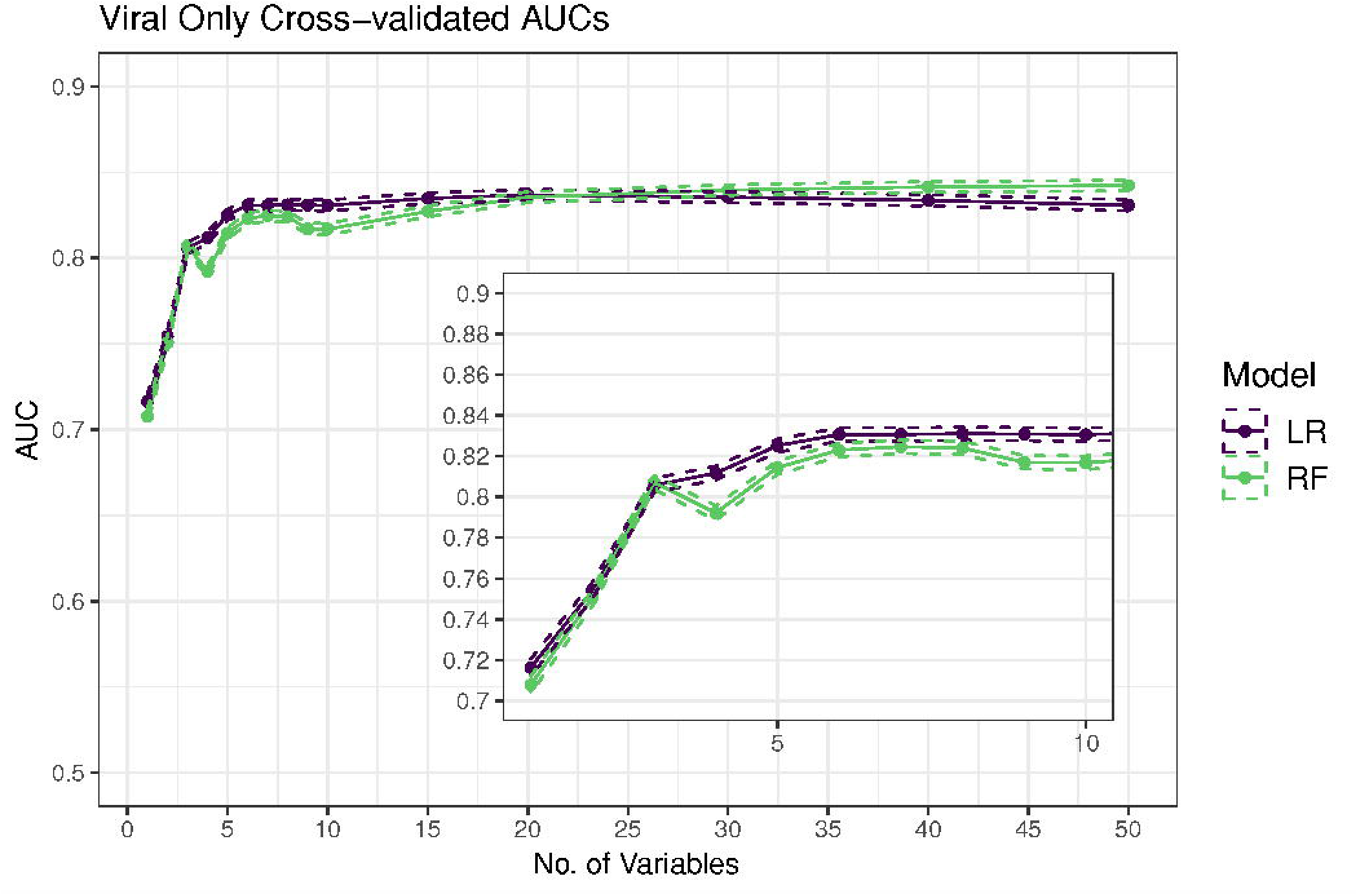
Average AUC and 95% CIs from cross-validation (100 iterations) for both a logistic regression (LR) and random forest (RF) as the number of variables in the model increases and inset shows zoomed in graphs of 1 through 10 variables.

When we examined the predictors associated with viral etiology for each of the 7 sites in GEMS by filtering the entire dataset by site, we found all had a similar order of variable importance with some minor differences (Table 4). We then looked at the predictions filtered for specific countries and specific continents within each cross-validation iteration’s test set to see how the model performs on these subgroups. We found that at Asian sites the predictions had an AUC almost 0.07 better than African sites on average. Looking at individual sites, in Kenya the model predictions had the worst average AUC while Bangladesh had the best average AUC. Across all sites, the AUC of a 5-variable model was similar to a 10-variable model with less than 0.02 lower average AUC.

**Table 4:**
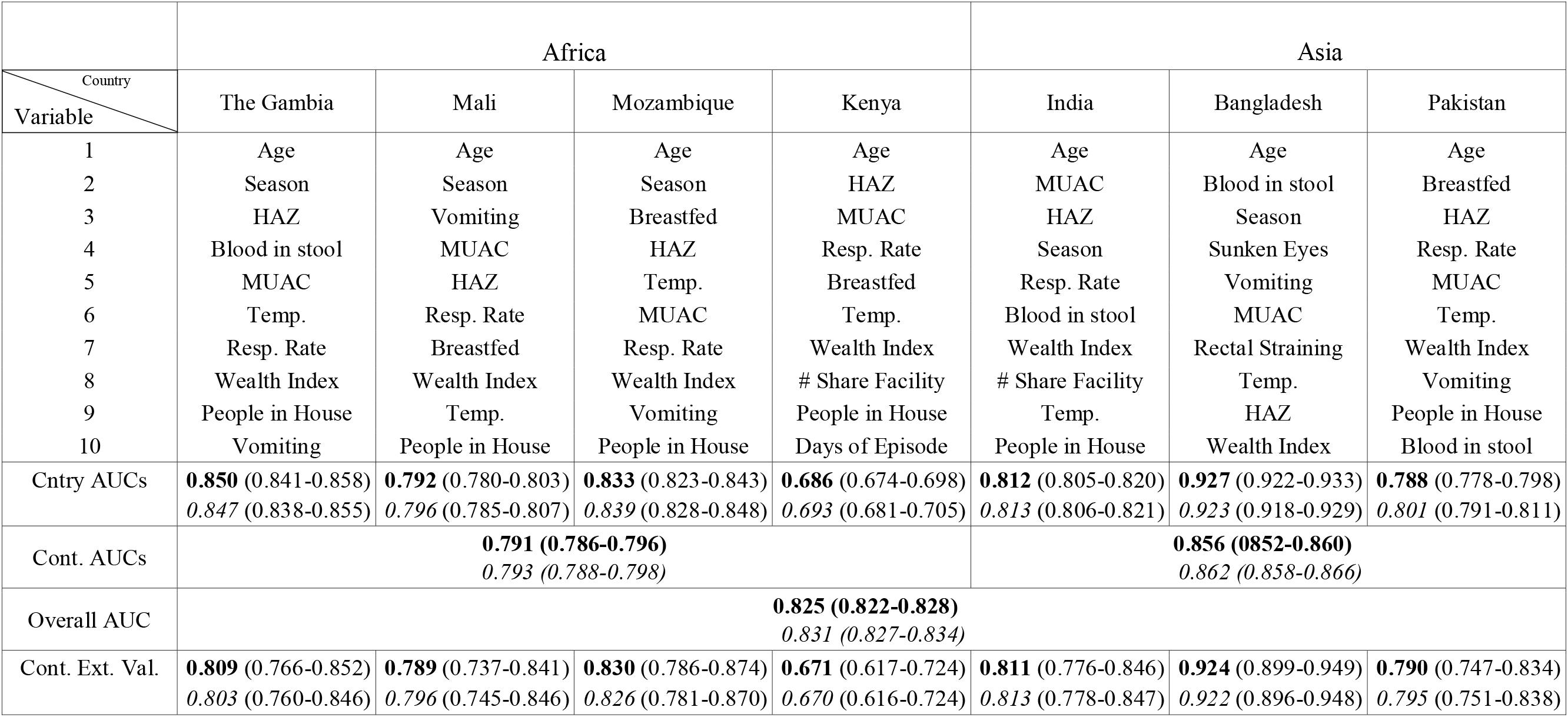
The table contains both site-specific variable importance ordering and a cross-validated average overall AUC, AUC by country, and AUC by continent and confidence intervals from a 5 (bold) and 10 (ital.) variable logistic regression model for predicting a viral etiology with variables based on the overall variable importance. Lastly, it shows the AUC and a 95% confidence interval resulting from testing the logistic regression with variables based on the overall variable importance on each site individually following its training on the other countries in the same continent

|  | Africa |  |  |  | Asia |  |  |
| --- | --- | --- | --- | --- | --- | --- | --- |
| Country<br>Variable | The Gambia | Mali | Mozambique | Kenya | India | Bangladesh | Pakistan |
| 1 | Age | Age | Age | Age | Age | Age | Age |
| 2 | Season | Season | Season | HAZ | MUAC | Blood in stool | Breastfed |
| 3 | HAZ | Vomiting | Breastfed | MUAC | HAZ | Season | HAZ |
| 4 | Blood in stool | MUAC | HAZ | Resp. Rate | Season | Sunken Eyes | Resp. Rate |
| 5 | MUAC | HAZ | Temp. | Breastfed | Resp. Rate | Vomiting | MUAC |
| 6 | Temp. | Resp. Rate | MUAC | Temp. | Blood in stool | MUAC | Temp. |
| 7 | Resp. Rate | Breastfed | Resp. Rate | Wealth Index | Wealth Index | Rectal Straining | Wealth Index |
| 8 | Wealth Index | Wealth Index | Wealth Index | # Share Facility | # Share Facility | Temp. | Vomiting |
| 9 | People in House | Temp. | Vomiting | People in House | Temp. | HAZ | People in House |
| 10 | Vomiting | People in House | People in House | Days of Episode | People in House | Wealth Index | Blood in stool |
| Cntry AUCs | **0.850** (0.841-0.858)<br>*0.847* (0.838-0.855) | **0.792** (0.780-0.803)<br>*0.796* (0.785-0.807) | **0.833** (0.823-0.843)<br>*0.839* (0.828-0.848) | **0.686** (0.674-0.698)<br>*0.693* (0.681-0.705) | **0.812** (0.805-0.820)<br>*0.813* (0.806-0.821) | **0.927** (0.922-0.933)<br>*0.923* (0.918-0.929) | **0.788** (0.778-0.798)<br>*0.801* (0.791-0.811) |
| Cont. AUCs | **0.791 (0.786-0.796)**<br>*0.793 (0.788-0.798)* |  |  |  | **0.856 (0.852-0.860)**<br>*0.862 (0.858-0.866)* |  |  |
| Overall AUC | **0.825 (0.822-0.828)**<br>*0.831 (0.827-0.834)* |  |  |  |  |  |  |
| Cont. Ext. Val. | **0.809** (0.766-0.852)<br>*0.803* (0.760-0.846) | **0.789** (0.737-0.841)<br>*0.796* (0.745-0.846) | **0.830** (0.786-0.874)<br>*0.826* (0.781-0.870) | **0.671** (0.617-0.724)<br>*0.670* (0.616-0.724) | **0.811** (0.776-0.846)<br>*0.813* (0.778-0.847) | **0.924** (0.899-0.949)<br>*0.922* (0.896-0.948) | **0.790** (0.747-0.834)<br>*0.795* (0.751-0.838) |

Given the logistic regression’s superior performance to random forest regression using 5 and 10 variables and in calibration, we performed validation by testing the logistic regression on each site individually following training on the other sites in the same continent, and found performance metrics similar to the cross-validation results, with AUC ranging from 0.65 to 0.92 across the seven sites. As with the internal cross-validation, we found 5-variable models to have similar performance to 10-variable models. We found similar results for the bacterial etiology prediction (Supplemental Table S8).

## Discussion

Our use of data from GEMS, which involved 3366 diarrheal episodes with known etiology in 7 countries and with over 150 clinically-relevant parameters collected for each episode, allowed for a robust analysis that revealed the ability of clinical variables alone to predict diarrheal etiology with a high degree of accuracy. Using machine learning algorithms, we found that a model with just 5 variables (age, season, HAZ, bloody diarrhea, and vomiting), could accurately predict viral etiology, with a cross-validated AUC of 0.825. Translation of these findings towards clinical decision making has the potential to improve management, including appropriate antibiotic use, in LMICs.

Previous studies predicting etiology of diarrheal illness [14-17], have been limited by the low number of participants, amount of clinical data collected, pathogen variety, number of pathogens detected, method of detection, lack of controls without diarrhea, single center design, and the need for stool testing. Etiological prediction is particularly challenging in LMIC settings, where multi-pathogen detection is common in children with diarrhea, and presumed pathogens can be isolated from asymptomatic individuals in up to 50% of study controls [18]. New molecular diagnostic methods used on the GEMS samples involved a quantitative assessment of 32 potential pathogens, with matched case-control pairs, to ascribe an etiological attributable fraction (AFe) for each episode. This quantitative method, in context of a case-control study, is thus able to account for the high rate of asymptomatic detection of pathogens by molecular testing in children in LMICs, which can confound the attribution of etiology. Using these data, we built several models to evaluate the effect of clinical indicators on whether children presenting with acute diarrhea had a viral etiology (or bacterial etiology). We showed that AUCs improved for the first 7 variables but thereafter the addition of more variables did not improve the model. Notably, we found that an AUC of *0.825* could be achieved with 5 variables, enabling the translation of this predictive model to a parsimonious rule which could be used in clinical decision-support. Additionally, we found that the random forest regression did not improve performance over regression models. This is likely due to the effect of the predictors on etiology being primarily linear. From the partial dependency plots, we show that, within the range of most of the data, the relationship between each predictor and the prediction is linear. Also, using interactions in a logistic regression model did not improve AUC.

When considering sensitivity and specificity in the context of diarrheal etiology, we assumed a high specificity target for prediction of “viral only” etiology (Figure 2), and similarly, a high sensitivity target for bacterial etiology (Figure S5), both of which would minimize the risk of not giving antibiotics to a child with a bacterial infection. While current WHO guidelines recommend antibiotics only for children with dysentery and for children with acute water diarrhea (AWD) with severe dehydration in cholera endemic regions, there is evidence suggesting treatment of non-dysenteric *Shigella* infections may be beneficial [19, 20]. Our prediction model showed that for predicting a viral etiology, for a desired specificity of 0.85, we achieved a sensitivity of 0.59. We found that the most significant predictors for differentiating viral from other etiologies were: age, HAZ, season, bloody diarrhea, and vomiting. Vomiting, a higher HAZ, and dry/cold season were evidence towards a viral etiology, while an older age and bloody diarrhea were evidence against a viral etiology.

**Figure 2:**
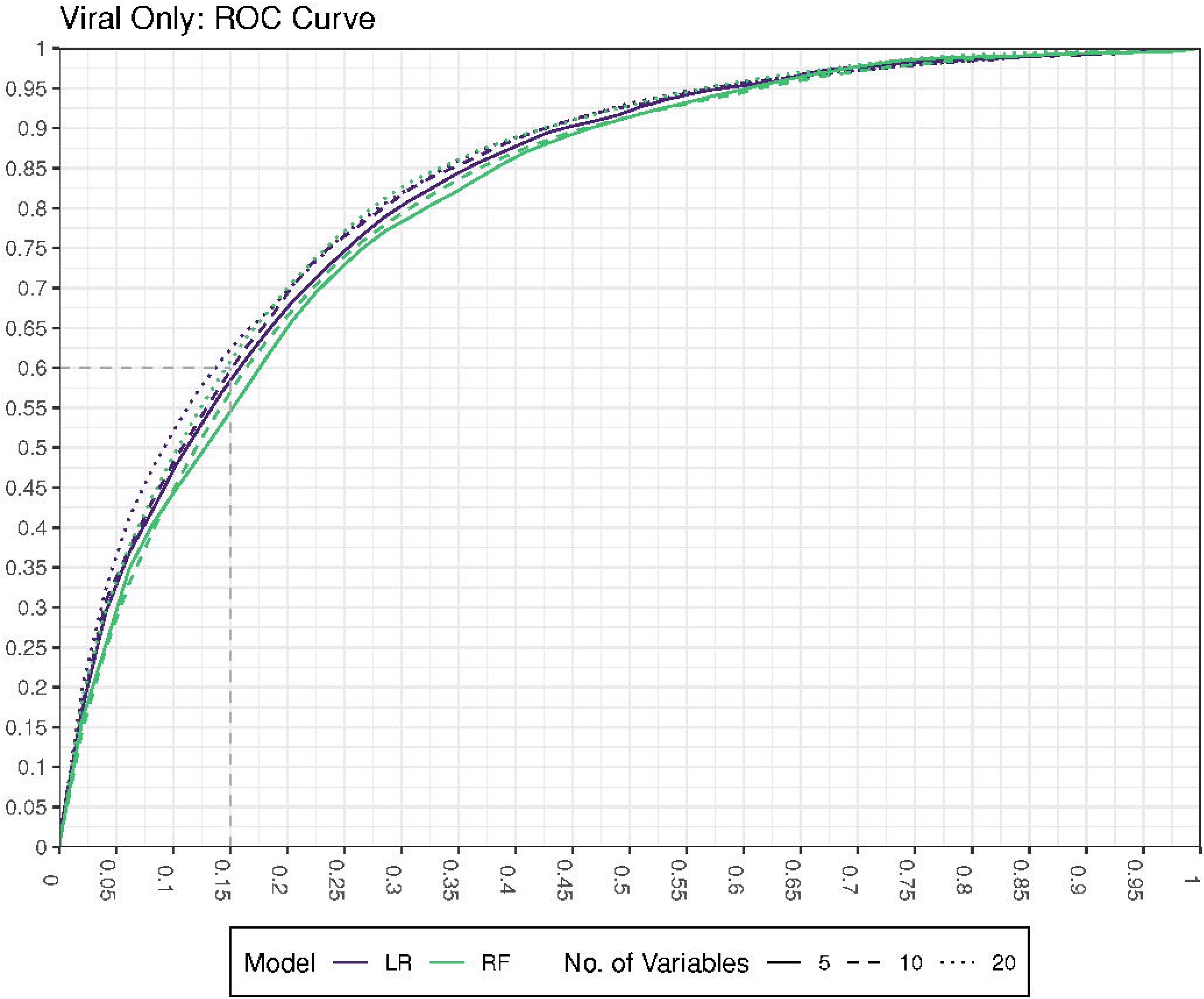
Interpolated estimates of ROC curves from the cross-validation for logistic regression and random forest models with variable sizes of 5, 10, and 20. The faded dashed lines represent examples of how we could achieve a sensitivity of 0.6 and a specificity of 0.85 for prediction of viral etiology.

The predictors we identified are consistent with those of previous studies. Bloody diarrhea as a predictor of a bacterial cause of diarrhea, especially for shigellosis, has been well established [14-17, 21-23]. and informs the IMCI guidelines that dysentery be treated with antibiotics. Vomiting as a predictor of a viral process has similarly been shown in previous studies [14, 16]. It is well established that younger children have a higher incidence of diarrhea [24] and some studies have suggested that younger age is also more indicative of a viral process [16, 22, 24-26].. We showed that age was the most important predictor with mean age of viral case being 13.0 months, and 22.1 months for bacterial cases.

Using data gathered from NOAA weather stations proximal to our study sites during the study period, we were able to develop seasonal variables based on temperature and rainfall. We show that a viral etiology of diarrhea is associated with a drier, colder climate, consistent with observation from previous studies from the USA [16] and India [26]. The positive association of anthropometrics (higher HAZ and mid-upper arm circumference (MUAC)) with viral etiology may suggest that improved nutrition is more protective of a bacterial than a viral process. Symptoms found in earlier studies to be predictive of etiology, but which did not improve predictive performance in our analysis when added to the variable importance selected variables include: fever, number of stools per day, duration of diarrhea, and presence of mucous [14-17, 23]. Similarly, variables related to hygiene and sanitation did not help with prediction of etiology.

Given that GEMS was conducted in 7 countries across Africa and Asia, we examined the model performance across sites. We found that the model attained an average AUC of about 0.86 in Asian sites and about 0.79 in African sites, likely due to poor performance of the model in Kenya and good performance in Bangladesh. This suggests that external validation will be necessary to assess both performance and generalizability. Indeed, even within continent, countries had varying AUCs. We also found that, when validated against other sites from the same continent by leaving one country out, use of five variables achieve similar AUC as use of 10 variables. Future studies should aim to capture country- or continent-specific trends such as background seasonality or sudden changes in climate or patient symptoms, so that outbreaks or volatility can be accounted for in the predictions.

Our study has a number of limitations. First, our predictive model does not distinguish between different bacterial etiologies or bacterial from parasite etiologies, which may require different therapy. Additionally, it does not predict for parasitic infections. In GEMS [6], a number of bacterial pathogens had few to no cases detected using AFe > 0.5, including EHEC, Yersinia, LT ETEC, EAEC, atypical EPEC, and Clostridium difficile. This was due to these organisms’ presence in control children without diarrhea, making attribution difficult. While it is possible that these could have co-occurred with a viral pathogen, there is limited evidence that antibiotic treatment of these etiologies would be beneficial in this setting. External validation is essential for this and all clinical prediction models, as demonstrated by our heterogenous result by continent. GEMS was conducted before the widespread use of rotavirus vaccine and rotavirus was the dominant viral pathogen; thus, the model will need to be validated in settings were rotavirus vaccination campaigns have had substantial impact. Although we present several measures of performance including sensitivity and specificity at various thresholds, we do not directly measure clinical usefulness. Future studies should explicitly show the potential for reduction in antibiotic use resulting from the clinical prediction. Lastly, our prediction models could be further adapted to individual clinical contexts, depending on the ease of obtaining different variables (i.e. availability of a height board versus a MUAC tape for anthropometric measurements).

In conclusion, utilizing a large number of cases and quantitative molecular methods of pathogen detection with etiologic attribution based on a case-control study, we showed that etiology prediction could be attained for episodes of acute diarrhea with as few as 5 variables. Our findings confirm previously considered predictors of viral etiology including lack of bloody diarrhea, vomiting, younger age, and a dry and cool climate, and reveal additional predictors of viral etiology associated with anthropometric measures. These findings have the potential to provide clinicians in lower-resource settings with better informed clinical decision making, including helping to identify a subset of children from whom antibiotics may be safely withheld and a group who may benefit from antimicrobials and/or adjunctive microbiologic testing.

## Data Availability

The full GEMS data is available at https://clinepidb.org/ce/app/ but you must apply for
permission to access the data. A prompt to request access opens when pressing the
download data button. Additionally, the website has a contact us page to provide a
point of contact.
The weather data can be downloaded freely at ftp://ftp.ncdc.noaa.gov/pub/data/noaa.

ftp://ftp.ncdc.noaa.gov/pub/data/noaa

## Supplementary Figure Legends

S1 Checklist: STROBE Checklist

Figure S2: The left map shows the locations of the 4 study sites in Africa. Right map shows the locations of 3 study sites in South Asia. The map was generated using the get_map and ggmap functions in R version 3.6.1.

Figure S3: Average AUC and 95% CIs from 100 iterations of cross-validation for both a logistic regression (LR) and random forest (RF) as the number of variables in the model increases and inset shows zoomed in graphs of 1 through 10 variables.

Figure S4: Consort diagram of the reduction of patients from 22567 in the GEMS dataset to the 3366 cases in our study. Note that we only filtered out non-responses for response variables that were in the top 50 of our screening step.

Figure S5: Average AUC and 95% CIs from cross-validation (100 iterations) for logistic regression (LR), random forest (RF), gradient boosted trees (GBR) and vanilla neural networks (NN) as the number of variables in the model increases and inset shows zoomed in graphs of 1 through 10 variables for just the top two models in this range, the LR and NN.

Figure S6: Partial dependency plots for the top ten important variable for a predicting a viral etiology. Ticks on the x-axis show the deciles of the data.

Figure S7: Histograms showing the distribution of predictions from the five variable model for both the patients with known etiologies and unknown etiologies determined by the greater than 0.5 AFe from TAC data.

Figure S8: Interpolated estimates of ROC curves from the cross-validation for logistic regression and random forest models with variable sizes of 5, 10, and 20. The faded dashed lines represent examples of how we could achieve a sensitivity of 0.85 and a specificity of 0.60 for any bacteria.

